# Patient safety education in undergraduate medical curricula: a concept analysis and scoping review protocol

**DOI:** 10.1101/2024.12.23.24318603

**Authors:** Jonathan Bowman-Newmark, Evangeline Brock, Helen Vosper

## Abstract

**Objective:** This study aims to establish how the phenomenon of patient safety education manifests within undergraduate medical curricula. The specific objectives are to conduct a concept analysis, in order to make explicit the concept of patient safety education and inform its operational definition. Thereafter, to conduct a scoping review, in order to systematically map what educational content is taught, who is taught that content, and how that content is taught.

**Introduction:** Preliminary searches identified four extant systematic reviews, published between 2010 and 2022, which investigated different aspects of how patient safety education manifested within undergraduate medical curricula. Notwithstanding, those findings do not provide a sufficiently robust basis to inform contemporaneous discourse.

**Eligibility criteria:** The participants, concept, and context framework will be used to determine the evidence sources eligible for inclusion in the scoping review. Eligible participants will include learners matriculated on undergraduate medical programmes, and the educators that teach learners on those programmes. The concept will be patient safety education, according to an operational definition that will be established. Eligible contexts will include the taught curricula of undergraduate programmes in medicine.

**Methods:** An exploratory and descriptive study that will sequentially implement the Walker and Avant framework for concept analysis and the updated JBI methodology for scoping reviews. Eight electronic databases and one internet search engine will be utilised to identify sources published between 1 May 2014 and 30 November 2024. Pre-defined eligibility criteria will be used to select sources. Data will be extracted and analysed, and findings will be presented.

## INTRODUCTION

Patient safety has been averred as “one of the most ancient and fundamental principles of medicine” (World Health Organization, 2024, p. vii); nonetheless, it still remains the case that many patients experience iatrogenic harm in healthcare settings. In an enduring paradox, safety tends to be signalled by its absence more than its presence (Reason, 2000; Ricciardi and Cascini, 2021). Plaintively, 6% of patients worldwide were estimated to have experienced an episode of preventable harm between 2000 and 2019 (Panagioti et al., 2019). Internationally, organisations including the General Medical Council and Medical Schools Council (2015)— respectively, the professional regulator for medical practitioners and the representative body for medical schools in the United Kingdom—the Association of American Medical Colleges (2024)—representative of accredited medical schools in North America—and the Government of India (2018) have all emphasised the importance of education as an intervention to reduce the future burden of harm. The importance of undergraduate patient safety education has been disputed though. Pedersen (2018) contended that “safety dispositions are not something that can be learnt and trained in the classroom” (p. 236), whilst conceding that “it is not unlikely that the seeds for the development of a pragmatic attitude can be planted in the classroom if the curriculum allows” (p. 236). Although it may be that “education alone is not the… answer” (Cosby, 2009, p. 282), patient safety education has been maintained as important in realising change (Al-Worafi, 2024), and ElAraby, Ra’oof and Alkhadragy (2018) even endorsed it as the “main solution to this problem” (p. 91).

Integration of patient safety education within healthcare professionals’ curricula has now been advocated for over two decades (Kohn, Corrigan and Donaldson, 2000; Association of American Medical Colleges, 2001; Institute of Medicine, 2001; Patient Safety Education Study Group, 2009; Lucian Leape Institute at the National Patient Safety Foundation, 2010). Indeed, international consensus exists that contemporary undergraduate medical curricula should include patient safety education (World Health Organization, 2009; 2011; 2021). Different educational strategies and modalities can be employed by educators to deliver patient safety education (Gilula and Barach, 2009), with teaching formats themselves being considered influential in facilitating effective learning (Maeda, Kamishiraki and Starkey, 2012). Moreover, making content representative of patient safety education ‘visible’ to learners has been judged particularly valuable (Abbott et al., 2020). Vincent and Amalberti (2016) emphasised how “labelling an issue as a safety issue is… strongly motivational” (p. 5), which reinforces Howe’s (2006) longstanding recommendation that at least “some part of a formal curriculum is explicitly focused on patient safety” (p. 188).

Preliminary searches, performed during October 2024, identified four systematic reviews that investigated different aspects of how patient safety education manifested within undergraduate medical curricula (Wong et al., 2010; Nie et al., 2011; Kirkman et al., 2015; Anugrahsari et al., 2022). Significantly, Nie et al. (2011) identified only a single previously-published report that described patient safety education within a core undergraduate medical curriculum. In turn, that report (Patey et al., 2007), described a literature review which did not identify any previous instances. Thus, it may be inferred that patient safety education—as a concept discussed in the literature in relation to undergraduate medical curricula—emerged only relatively recently. In 2009, Sandars posed the questions: ‘what’ content should be learned, and ‘how’ should that content be taught? The same year, the World Health Organization provoked consideration of ‘when’ patient safety education should be taught in their publication the ‘WHO Patient Safety Curriculum Guide for Medical Schools’. Kirkman and colleagues’ (2015) systematic review, which updated the earliest of the four systematic reviews of patient safety education (Wong et al., 2010), provided robust and contemporaneous insights. Nevertheless, its findings were derived from searches that are now more than a decade old. Patient safety advocates have more recently highlighted the “massive efforts underway to strengthen patient safety education in medical schools” (Watson, 2017, p. 12), which Pedersen (2018) concurred was “increasingly becoming an obligatory part of the curriculum in medical schools worldwide” (p. 233). Moreover, Leape (2021), who echoed Gandhi et al. (2018), intimated that undergraduate medical “curricula have increasingly included patient safety and safety science” (p. 376).

Notwithstanding the methodological limitations of the most contemporaneous systematic review available (Anugrahsari et al., 2022), those perspectives would appear substantiated. However, weaknesses in respect of that study’s conduct and reporting constrain its relative contribution, and in isolation the study does not provide a sufficiently robust basis to inform contemporaneous discourse. Forward citation searches, performed during October 2024, of the known systematic reviews did not identify more recent systematic studies that updated their findings. This suggests that an evidence synthesis to address a widening gap in the literature would now be timely.

It was conspicuous, across the preliminary literature reviewed, that patient safety education was conceptualised inconsistently through use of various terminologies. This may lead to a potential conflation of two closely-related but distinct concepts: safety and quality. Although safety scientists have predicated safety in terms of quality (Hollnagel, 2014; Hollnagel, Wears and Braithwaite, 2015), different perspectives have been contended as to “whether one is a subset of the other and, if so, which [is] superordinate” (Wears and Sutcliffe, 2019, p. 42). Vincent (2010) intimated that many are content to simply describe their relationship “as a continuum” (p. 37). Perhaps in an implicit appreciation of that, Wong et al. (2010) and Kirkman et al. (2015) opted to examine patient safety and quality improvement education concurrently. Weingart’s (2005) critique that “the terminology of safety is perplexing on a good day, and near impossible on a bad one” (p. 93) seems pertinent here. Although “the perimeter of safety” has expanded (Vincent and Amalberti, 2015, p. 539), it has long been acknowledged that the dimensions of safety and quality “are not synonymous” (Cooper et al., 2000, p. 3). Accordingly, it seems important for any future research to unambiguously delineate the meaning of patient safety education, in order that ensuant findings are specific to the concept of interest.

Concept analysis is a rigorous method of examining a concept and its use in order to ascertain its attributes (Delves-Yates, Stockl and Moore, 2018). Significantly, concept analysis has been considered particularly suited to analysing emerging concepts that arise from practice (Yazdani and Shokooh, 2018). Walker and Avant’s (1983; 2019) linguistic framework has been recognised as the predominant approach to concept analysis used by researchers (Rodgers, Jacelon and Knafl, 2018). Furthermore, it has been deemed the “most comprehensive and systematic method” (Chabeli, Nolte and Ndawo, 2021, p. 13). A schematic representation of this framework is shown (Figure 1).

**Figure 1.**
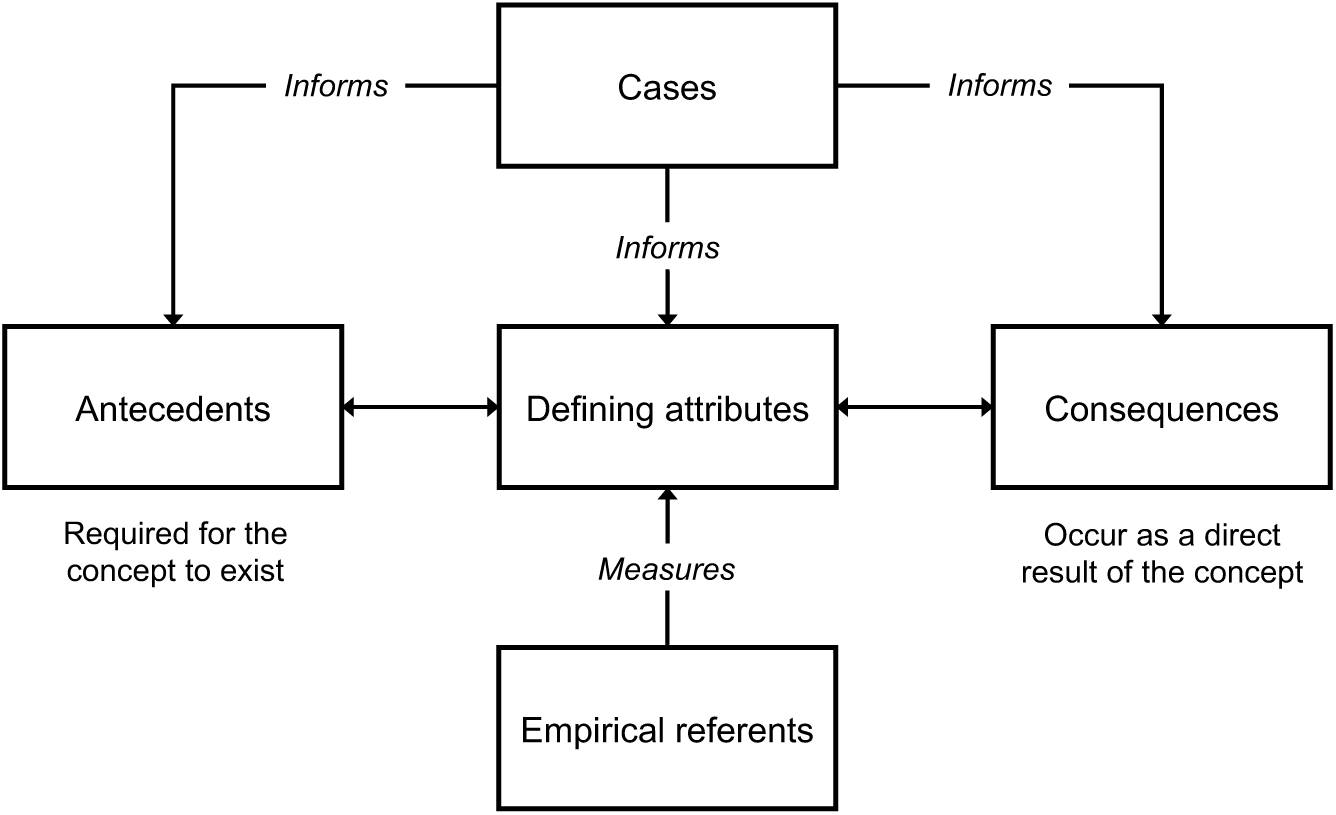
Schematic representation of the concept analysis framework (adapted from Walker and Avant, 2019, p. 180)

Walker and Avant (2019) acknowledged this framework’s utility in analysing a concept “objectively as subject matter, rather than subjectively as a persuasive weapon” (p. 183), which may bound out known tendencies to ‘moralise’ when a concept has value implications. It is expected this may be propitious when examining the concept of patient safety education, given its implicit and explicit meanings. Although criticism has been offered that concept analysis can be over-reductionist with respect to complex concepts (Weaver and Mitcham, 2008), it is proposed here that an operational definition of patient safety education—informed through concept analysis—will afford opportunities to facilitate more robust evidence synthesis processes. Explicitly defining the concept of interest is expected to positively impact on subsequent source selection, and data extraction and analysis. Preliminary searches, performed during September 2024, did not identify a previously-published concept analysis of patient safety education. Undertaking this now, in order to inform a properly-situated understanding (Toulmin, 1972; Bergdahl and Berterö, 2016), is proposed as a means to objectively operationalise its definition (Waltz, Strickland and Lenz, 2017; Grove and Gray, 2023). That approach remains entirely consonant with the perspective that concept analysis “should be a bridge to further related work” (Walker and Avant, 2019, p. 181), and partially rebuts Draper’s (2014) criticism that concept analysis makes an insubstantial contribution to scholarship.

In 2015, Wong offered a prospective research agenda that outlined specific patient safety education research needed as this related to medical curricula. Despite Zhou and colleagues’ (2024) demonstration of growing trends in patient safety education research over recent years, Wong’s agenda still remains apposite today. Moreover, it remains undisputed that “medical curricula must be challenged and changed to educate the clinicians that we need in the future” (Lachman, 2021, p. 48); as a consequence, robust evidence is continually needed to inform that change. Accordingly, a systematic evidence synthesis aimed at understanding how the phenomenon of patient safety education manifests within undergraduate medical curricula would satisfy both the expressed and perceived needs of knowledge users (Pollock et al., 2022). However, it is proposed here that such an evidence synthesis should not seek only to update the existing systematic reviews. In common with expectations arising from other relatively-novel concepts of interest, it is anticipated that the concept of patient safety education has evolved as knowledge has been accrued (Perneger, 2006; Chinn, Kramer and Sitzman, 2022). Instead, a broader exploratory research question is proposed that would more optimally be addressed through a scoping review (Munn et al., 2018; 2023; Peters et al., 2021; Pollock et al., 2024). Utilisation of a scoping review design in these circumstances is supported by the Knowledge Translation Program’s ‘Right Review’ decision support tool (Amog et al., 2022; Clyne et al., 2024). Preliminary searches of the Open Science Framework registries and International Prospective Register of Systematic Reviews, performed during October 2024, did not identify any similar study that was ongoing or recently completed.

Therefore, this study aims to establish how the phenomenon of patient safety education manifests within undergraduate medical curricula. The specific objectives are:

1. to conduct a concept analysis, in order to make explicit the concept of patient safety education and inform its operational definition; thereafter,
2. to conduct a scoping review, in order to systematically map what educational content is taught, who is taught that content, and how that content is taught.

## REVIEW QUESTIONS

This study aims to answer the primary review question: how does the phenomenon of patient safety education manifest within undergraduate medical curricula? That question will be addressed by synthesising answers to three sub-questions that seek to explore the concept with respect to the study’s participants and context:

1. what content representative of patient safety education is included within undergraduate medical curricula?
2. which undergraduate medical student cohorts are the learners of that content?
3. which pedagogic approaches are used by educators to deliver that content?

## ELIGIBILITY CRITERIA

The participants, concept, and context framework will be used to determine the evidence sources eligible for inclusion in the scoping review.

### Participants

Eligible participants will include learners matriculated on undergraduate educational programmes in medicine, and the educators that teach learners on those programmes.

### Concept

The concept of interest will be patient safety education, expressed and understood according to an operational definition that will be established.

### Context

Eligible contexts will include the taught curricula of undergraduate programmes in medicine. Taught curricula comprise “the content that is delivered through educational activities for the students to learn from” (Chakrabarti et al., 2021, p. 2) and reifies that which occurs in practice (Wu, 2022; Seifouri, 2023). Any interprofessional educational contexts, where these are reported, must identifiably include eligible participants. There will be no restrictions imposed with respect to geographical location.

### Types of evidence sources

The scoping review will consider undifferentiated, published or unpublished, peer-reviewed or non-peer-reviewed primary research of any quantitative, qualitative, or mixed-methods methodology. Anticipated study designs include, but will not be limited to, experimental, quasi-experimental, analytical and descriptive observational, ethnographic, and qualitative description. Evidence syntheses, including systematic reviews and scoping reviews, will also be considered. Sources with titles and abstracts not published in English, and / or retracted post-publication, will be excluded.

## METHODS

This exploratory and descriptive study will implement concept analysis and scoping review methods sequentially, in a manner analogous to the novel methodology described by Kang, Kim and Cho (2023).

### Concept analysis

A concept analysis of patient safety education will be conducted according to the eight-step framework described by Walker and Avant (1983; 2019), which itself represents a modified version of Wilson’s (1963) classical analysis procedure:

1. select the concept (viz. patient safety education);
2. determine the aims of analysis (i.e., to inform an operational definition);
3. identify uses of the concept;
4. determine its defining attributes;
5. identify a model case (i.e., a paradigmatic exemplar);
6. identify additional cases (e.g., borderline, related, and contrary cases);
7. identify antecedents and consequences; and
8. determine empirical referents (i.e., tangible and measurable phenomena).

As illustrated in Figure 1, Walker and Avant’s framework necessitates iterative processes (Yazdani and Bayazidi, 2020). Despite each step being numbered sequentially, movement between steps is expected.

### Scoping review

The scoping review will be conducted in accordance with the updated JBI methodology (Peters et al., 2020a; 2020b), which itself is based on Arksey and O’Malley’s (2005) foundational framework as enhanced by Levac, Colquhoun and O’Brien (2010). The study will be reported in accordance with the preferred reporting items for systematic reviews and meta-analyses (PRISMA) extension for scoping reviews (PRISMA-ScR) (Tricco et al., 2018), with the proviso that this guidance is currently being updated (Tricco et al., 2024). A completed PRISMA-ScR checklist will be made available as an appendix to the final report. This a priori protocol was developed in accordance with best practice guidance and the reporting items for scoping review protocols (Peters et al., 2022). The proposed study was prospectively registered with the Open Science Framework (https://doi.org/10.17605/OSF.IO/Z9VQ7).

#### Search strategy

The search strategy is intended to systematically and comprehensively identify potentially-relevant sources, within the constraints of available resources and time. Electronic bibliographic databases and an internet search engine will be utilised. In order to identify sources available following Kirkman and colleagues’ (2015) systematic review, searches will target sources published between 1 May 2014 and 30 November 2024, inclusive.

Firstly, in order to identify potentially-relevant seed sources, initial searches of MEDLINE (Ovid) and Embase (Ovid) databases will be performed using search facets based on a logic grid aligned to the eligibility criteria. This is summarised as a draft search strategy (Appendix A). Secondly, text words contained in the titles and / or abstracts of potentially-relevant seed sources, and the controlled vocabulary used to index those sources, will be used to develop a primary search strategy for MEDLINE (Ovid). Guidance will be sought from an information specialist to optimise the specificity of the search, in order to manage the volume of citations whilst realising a desire for sensitivity (Peters et al., 2020b; Alexander et al., 2024). In order to quality assure its integrity the search strategy will be peer reviewed by an information specialist not otherwise associated with this study, in line with the peer review of electronic search strategies (PRESS) guideline (McGowan et al., 2016). The peer-reviewed primary search strategy will be made available as an appendix to the final report. To negotiate expected variability in indexing between bibliographic databases, and to limit the impact of publication biases, the primary search strategy will then be adapted for five additional biomedical or interdisciplinary electronic databases: Embase (Ovid), CINAHL (EBSCO), Cochrane Library, PsycInfo (Ovid), and Web of Science (Clarivate). As recommended by Lam et al. (2024), ERIC—an education-specific electronic database—will also be searched using EBSCO. Wherever possible, translation of the primary search strategy’s search strings will be technology augmented in order to better align this technical task with recognised human capacities and capabilities (O’Connor et al., 2019; Khalil et al., 2024). Polyglot Search Translator will be used, which is validated for this purpose (Clark et al., 2020). In order to constructively address a potential lack of inclusivity resulting from publication biases, grey literature will be searched using Dissertations & Theses and A&I (ProQuest) and one internet search engine (Google; first five pages of search hits) (Pearson et al., 2024). Thus, any consequent exclusion of potentially-relevant sources can be acknowledged and rationalised (Peters et al., 2022). In order to ensure any retractions are identified, searches will seek to capture post-publication amendments (Lefebvre et al., 2024). Thirdly, the reference lists of included sources will be hand searched for additional potentially-relevant sources.

All identified citations will be collated using reference management software (RefWorks; ProQuest, Ann Arbor (MI), United States of America). Citations will then be de-duplicated using Systematic Review Accelerator Deduplicator (Forbes et al. 2024), which has demonstrated superior accuracy and sensitivity for automated de-duplication (McKeown and Mir, 2024). If sources are found to be replicated then the most up-to-date version of the source will be preferred.

#### Source of evidence selection

Eligibility criteria will be used to select sources according to a two-stage process, which will involve the screening of titles and abstracts followed by the screening of full texts. Following collaborative development of an elaboration document, intended to support objective source selection, two human reviewers (JB-N and HV) will independently conduct screening processes in duplicate using Rayyan (Cambridge (MA), United States of America). Any disagreements that arise with respect to eligibility will be resolved through discussion. Should consensus not be reached then disagreements will be arbitrated by a third human reviewer (EB).

Although not yet ascertained empirically, it has been suggested that the corpus of literature that may require to be screened could be extensive on account of the continued emergence of the concept of interest (Wong, 2015; Zhou et al., 2024). Should the volume of potentially-relevant sources prove too extensive to be practicable following title and abstract screening, decisions about restricting the number of full text sources screened will prioritise state-of-the-art (Alexander et al., 2024). In order to answer the same review questions, albeit for a narrower, more contemporaneous time interval, newer evidence sources will be preferred.

Following the screening of titles and abstracts, potentially-relevant sources whose full text is presented in a language other than English will be excluded. Resource constraints preclude their professional translation. No-cost neural translation will not be performed due to an inability to confirm the interpretation of a nuanced concept of interest, which is expected to be determined in part by “cultural, contextual, and societal differences” (Kim et al., 2015). However, in order to manage potential reporting biases, the number of citations involved will be reported alongside other sources not included following full text screening. A list of the citations involved will also be made available as an appendix to the final report. Sources that appear to satisfy eligibity criteria, or where uncertainty still remains, will be retrieved using institutional access policies or requested as inter-library loans. Where a source cannot be obtained via those means, the source’s corresponding author will be contacted by e-mail—maximum of two attempts—to seek the source prior to its exclusion. To facilitate the screening of full texts, all retrieved full texts will be imported to Rayyan. Where uncertainty remains about a source’s eligibility following full text screening, the source’s corresponding author will be contacted by e-mail—maximum of two attempts—in an attempt to resolve question(s) prior to its exclusion. Identical processes will be followed for any potentially-relevant sources identified from the reference lists of included sources.

A preliminary pilot screening exercise will be conducted prior to the screening of titles and abstracts, which will involve the intended reviewers examining a proportion of randomly selected sources. This will occur again prior to the screening of full texts. The reviewers’ inter-rater agreement will be calculated and will require to be ≥75% before each stage, in accordance with available best practice guidance for scoping review conduct (Peters et al., 2020b). If the target is not achieved initially, the reviewers will discuss their reasons for agreement and disagreement; reviewers will undertake repeated cycles of piloting and collaborative modification of the source selection elaboration document until the target is achieved.

The number of sources identified, duplicates removed, sources screened, and sources excluded following full text screening, alongside the reasons for exclusion, will be reported narratively and as a PRISMA flow diagram (Page et al., 2021).

#### Data extraction

A standardised electronic data extraction instrument will be developed as a workbook in Microsoft Excel for Mac version 16.78 (Redmond (WA), United States of America), based on a draft data extraction instrument (Appendix B). The data items extracted will be guided by the review sub-questions and variables that have been reported previously (Wong et al., 2010; Nie et al., 2011; Kirkman et al., 2015). A data extraction elaboration worksheet will be developed to promote consistency during data extraction. The variables extracted are expected to evolve as data extraction progresses; as such, this worksheet will become a ‘living’ document. In order to ensure that data is reliably captured, a preliminary pilot data extraction exercise will be conducted by the intended data extractor (JB-N) using a selection of different source types. Any missing, redundant, or unclear variables will be added, removed, or revised. Where additional data is required, a source’s corresponding author will be contacted by e-mail—maximum of two attempts—on a case-by-case basis. Due to resource constraints, data extraction will be undertaken by a single data extractor only. The final data extraction instrument will be made available as an appendix to the final report.

A planned simplification is proposed with respect to the extraction of content representative of patient safety education. The World Health Organization’s (2011) patient safety topics (pp. 18–19)—incorporating themes predicated on an existing evidence-based education framework (Australian Council for Safety and Quality in Health Care, 2005; Walton et al., 2006)—will be utilised to guide categorisation. In order to promote increased consistency during data extraction, the data extraction instrument will incorporate a closed-ended 11-item pre-populated matrix intended to operationalise structured data collection as a preliminary stage of data cleaning.

Following its extraction, the raw data will be cleaned by the data extractor. In order to manage potential ascertainment biases, a proportion of the data will be checked by one verifier (HV) for accuracy and completeness (Robson et al., 2019). Any disagreements that arise about the data extracted will be resolved through discussion. Should consensus not be reached then disagreements will be arbitrated by a third party (EB). Data extraction processes undertaken for other systematic forms of evidence synthesis have been reported not to be significantly affected by a data extractor’s level of experience (Jones et al., 2005; Horton et al., 2010). Accordingly, pragmatic decision making about the utilisation of resources during this study will prioritise multiple researchers’ involvement in screening processes, in order to mitigate against a possibly greater risk that potentially-relevant resources may be excluded inappropriately following single-reviewer screening (Waffenschmidt et al., 2019; Gartlehner et al., 2020).

#### Data analysis and presentation

Contextualised descriptive analyses will synthesise how the phenomenon of patient safety education manifests within undergraduate medical curricula. On account of the anticipated heterogeneity of sources, the review sub-questions themselves will comprise the principal units of analysis. No formal appraisal of methodological quality or risk of bias will be undertaken, and evidence synthesis will focus on high-level categorisation. Those approaches remain entirely consonant with available best practice guidance for scoping review conduct (Peters et al., 2020a; 2020b; Pollock et al., 2023).

Characteristics of included sources and review findings will be presented in narrative, tabular, and / or visual formats appropriate for scholarly discourse (Nyanchoka et al., 2019). Their rendering will be progressively refined to maximise clarity of presentation, and will proactively anticipate how to engage knowledge users (Khalil et al., 2021; Pollock et al., 2023). Potential visual formats, which may be created using either proprietary or open-source software, may include: a word cloud, to aggregate the content included within undergraduate medical curricula representative of patient safety education; a waffle chart, to differentiate undergraduate medical student cohorts as the learners of that content; and / or iconography, to illustrate pedagogic approaches used to deliver that content.

## DISSEMINATION

The findings of this study will be submitted for publication in a high-impact, discipline-specific peer-reviewed scholarly journal. It is anticipated that those findings will be likely to impact at an academic level; consequently, findings may also be disseminated through academic conference presentations. To promote engagement with prospective knowledge users, a short video abstract will also be produced.

## DEVIATIONS FROM PROTOCOL

On account of the iterative processes proposed—particularly execution of the search strategy, data extraction, and data analysis and presentation—deviations from this protocol may be required. Significant deviations will be tracked using a worksheet in Microsoft Excel for Mac version 16.78 (Redmond (WA), United States of America), and these will be reported and explained in the methods of the final report. That approach remains congruent with available best practice guidance for reporting deviations from a scoping review protocol (Peters et al., 2022).

## ACRONYMS

PRESS: peer review of electronic search strategies
PRISMA: preferred reporting items for systematic reviews and meta-analyses
PRISMA-ScR: preferred reporting items for systematic reviews and meta-analyses extension for scoping reviews

## ACKNOWLEDGEMENTS

The authors wish to thank Mel Bickerton (Information Consultant and Site Services Manager, Medical Library, University of Aberdeen) who provided guidance in the development of the draft search strategy.

## ETHICS

Ethical approval is not required for this study, which is a concept analysis and scoping review of previously-published data otherwise available in the public domain.

## DATA AVAILABILITY STATEMENT

Data sharing is not applicable to this protocol as no new data were created or analysed.

## CRediT AUTHOR CONTRIBUTIONS

Conceptualisation: JB-N and HV; data curation: JB-N; investigation: JB-N, EB and HV; methodology: JB-N, EB and HV; project administration: JB-N; resources: JB-N; supervision: EB and HV; visualisation: JB-N; writing—original draft: JB-N; writing—review and editing: JB-N, EB and HV. JB-N is the guarantor of this protocol.

---

**APPENDIX A.**
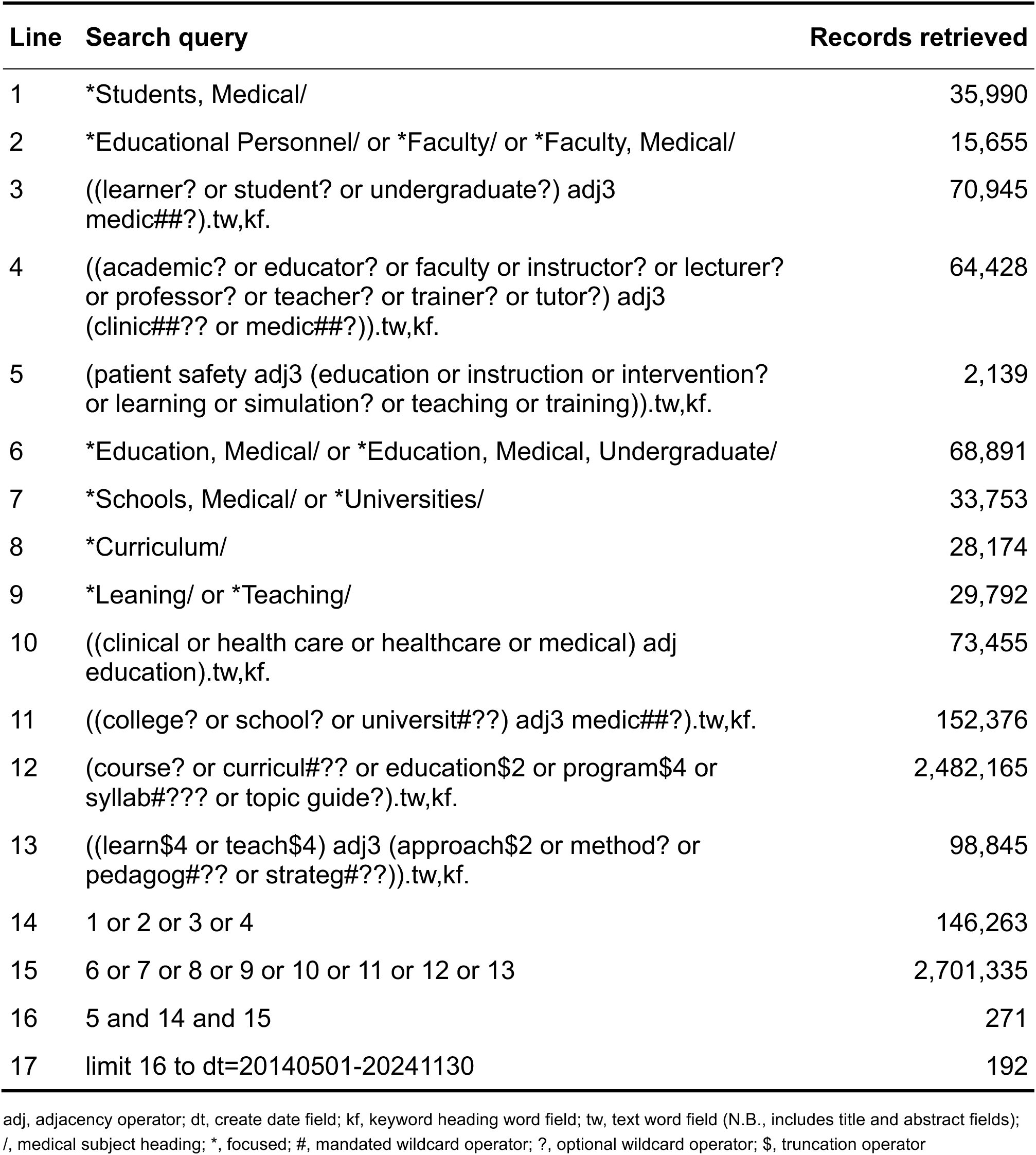
Draft search strategy. Search of MEDLINE (Ovid) performed on 3 December 2024

**APPENDIX B.** Draft data extraction instrument.

| Source citation and research design |  |  |  | Contextual characteristics |  |  | Key findings with respect to the review sub-questions |  |  |  |  |  |  |  |  |  |  |  |  |
| --- | --- | --- | --- | --- | --- | --- | --- | --- | --- | --- | --- | --- | --- | --- | --- | --- | --- | --- | --- |
| First author | Year | Citation details (e.g., journal; volume(issue), page(s)) | Research design | Medical school / university (Country) | Description of the curriculum | Curriculum implementation (i.e., year–year) | What content representative of patient safety education is included within the curriculum? <sup>a</sup> |  |  |  |  |  |  |  |  |  |  | Which medical student cohorts are the learners of that content? | What pedagogic approaches are used by educators to teach that content? |
|  |  |  |  |  |  |  | 1 | 2 | 3 | 4 | 5 | 6 | 7 | 8 | 9 | 10 | 11 |  |  |
| First author | 20XX | Journal; XX(X), XX–XX | Description of the research design | Medical school / university (Country) | Description of curriculum | XXXX–XXXX | x | x | x | x | x | x | x | x | x | x | x | Description of the medical student cohort(s) | Description of the pedagogic approach(es) used |

- Legend - 1. What is patient safety? - 2. Why applying human factors is important for patient safety - 3. Understanding systems and the effect of complexity on patient care - 4. Being an effective team player - 5. Learning from errors to prevent harm - 6. Understanding and managing clinical risk - 7. Using quality improvement methods to improve care - 8. Engaging with patients and carers - 9. Infection prevention and control - 10. Patient safety and invasive procedures - 11. Improving medication safety
<sup>a</sup> A closed-ended pre-populated matrix is proposed to categorise content representative of patient safety education. The 11 items correspond to the World Health Organization’s (2011) patient safety topics (pp. 18–19).

